# Domains included in multidimensional sleep health composite scores: A scoping review

**DOI:** 10.1101/2025.05.06.25327078

**Authors:** Catherine Siengsukon, Jade Robichaud, Ashley Barry, Prasanna Vaduvathiriyan, Karen Bock

## Abstract

While the SATED/Ru-SATED sleep health framework is well-recognized, there is no consensus for how many or which domains should be included in the construct of a multidimensional sleep health (MDSH) composite score or how the domains are defined. Therefore, the purpose of this scoping review is determine what domains are included in MDSH composite scores and how are those domains assessed. The Preferred Reporting Items for Systematic Reviews and Meta-Analyses extension for scoping reviews (PRISMA – ScR) and the Joanna Briggs Institute’s (JBI) updated methodology for scoping reviews were used. Two authors independently reviewed titles/abstracts, full-texts, and performed data extraction, and one author resolved any discrepancies with discussion as needed. The search strategy generated 1,481 references, 107 underwent a full-text screening, and 39 were eligible for inclusion in this scoping review. Five and six domains were the most common number of domains included in the MDSH composite scores (n=17 for both). Seventeen articles included only self-report measures and n=19 articles included a mix of self-report and objective measures. Thirteen unique domains were identified with Duration being the most common (n=38), followed by Alertness/Sleepiness (n=35), Satisfaction/Quality (n=32), and Timing (n=31). In conclusion, while the domains most often included in the MDSH composite scores followed the SATED/Ru-SATED framework, there was variability in the domains included as well as variability in how the domains were assessed. Consensus is needed on the definition of sleep health domains or, at a minimum, clear reporting on the definitions used. Further research is needed to determine which MDSH domains are most associated with health outcomes.

## 1. INTRODUCTION

Sleep medicine has historically focused on the identification and treatment of sleep disorders, such as insomnia and sleep apnea. *Sleep health* is a newer term that focuses on sleep as a multidimensional health behavior along a continuum with excellent sleep health on one end of the continuum and very poor sleep health on the other end, and sleep health exists in the presence *or absence* of a sleep disorder. For example, chronic insomnia occurs in approximately 10% of the US population,^1^ whereas approximately 60% of adults have short sleep duration (< 7 hours).^2^ Also, about 50% of adults report not feeling well-rested, and approximately 70% have inconsistent bedtime and waketime.^3^ Importantly, various dimensions of sleep health have been associated with adverse health consequences^4^, including increased risk of Alzheimer’s Disease (AD)^5-8^ and decline in cognitive function.^9^

When first introduced as a construct^4^, sleep health was proposed to consist of five domains: <u>S</u>atisfaction with sleep, daytime <u>A</u>lertness, <u>T</u>iming of sleep within 24 hour period, sleep <u>E</u>fficiency, and sleep <u>D</u>uration (acronym SATED). Regularity was added as a proposed domain shortly thereafter (acronym Ru-SATED).^10^ However, there is no consensus for how many or which domains should be included in the construct of sleep health.^11^

Because sleep health is a multidimensional construct, it is not intended to be measured by a single domain (such as sleep quality) but rather a combination of multiple domains, frequently reported as a multidimensional sleep health (MDSH) composite score. While the Ru-SATED sleep health framework is perhaps the most recognized framework, there is no consensus on the domains included in a MDSH composite score or the sleep characteristics used to define the domains.^11^ In addition, the sleep characteristics included in each domain can be measured in different ways (i.e. self-report, actigraphy, polysomnography).^11^ This has resulted in variable MDSH constructs and scoring techniques that make it difficult to compare research results. Therefore, the objective of this scoping review is to answer the questions, “ What are the domains included in multidimensional sleep health (MDSH) composite scores and how are those domains assessed?”

## 2. METHODS

This scoping review was reported as per the Preferred Reporting Items for Systematic Reviews and Meta-Analyses extension for scoping reviews (PRISMA – ScR)^12^ standard and the Joanna Briggs Institute’s (JBI) updated methodology for scoping reviews.^13^ A protocol was developed and registered in the Open Science Forum (https://osf.io/zhfya) for conducting this scoping review.^14^

The research librarian (PV) developed a pilot search strategy on the following databases: Ovid MEDLINE(R) and Epub Ahead of Print, In-Process, In-Data-Review & Other Non-Indexed Citations, Daily and Versions <1946 to June 17, 2024>. To construct the search strategy, the key concept identified was sleep health as the phenomena of interest and multidimensional measures of sleep as the measurement properties. The pilot search was completed on June 18^th^, 2024. Additional resources including PsycINFO, Embase, and Web of Science were searched along with selected cited references search and grey literature. The complete search strategy is available in Supplementary File 1.

Criteria for study inclusion were: 1. Included human participants, 2. Included ≥ 4 sleep health domains in a multidimensional sleep health composite score (the study did not have to use the term “ sleep health” to describe the composite but the term used must be in the spirit of sleep health; studies that assessed domains separately but did not calculate a composite were not included; the study was included if they defined the domains to be included in the composite a priori), and 3. Utilized an observational or interventional study design assessing sleep health through a composite score. Studies were excluded if they: 1. Included animals, 2. Were written in a language other than English, 3. Were published prior to 2014, 4. Data was published as an abstract or in non-peer reviewed sources (i.e. conference proceedings, preprint), 5. Were irrelevant to the study question.

Citations generated from the search strategy were exported to EndNote (version 21), and duplicates were removed. Citations were then imported to Covidence (Veritas Health Innovation, Melbourne, Australia; www.covidence.org) to review and perform data extraction. Each article title and abstract was independently reviewed by two reviewers (AB and KB) to determine eligibility based on the inclusion/exclusion criteria, and discrepancies were resolved by a third reviewer (CS). The full-text articles were then reviewed independently by two of four reviewers (AB, KB, JR, or CS) to determine eligibility, and discrepancies were resolved by CS.

A standard data extraction form was built in Covidence by CS. All study personnel involved in data extraction reviewed and approved the data extraction form prior to the start of data extraction. Data was independently extracted from eligible articles by two of four reviewers (AB, KB, JR, or CS). CS then reviewed the data extraction for accuracy, completeness, and to reach consensus. Extracted data was reviewed for accuracy by the primary reviewer, and a third reviewer was used to resolve discrepancies in data when needed. Data extracted included last name of first author, publication year, country study was conducted, study purpose/aim, the name of each sleep health domain, if each domain was assessed using self-report or objective means, how each domain was assessed (ie. question or questionnaire used, objective device used), possible responses for each domain, how each domain was defined as “ good” sleep, how the sleep health composite was calculated, description of the MDSH scale, details about the study sample (ie. pertinent comorbid conditions, targeted age group, location participants living/residing, same size, age, ethnicity, race, and sex), and main sleep health results.

Descriptive statistics (i.e. number and percentage) were calculated, including the number of domains included in the MDSH composite score and if self-report only, objective measures only, or mix of self-report and objective measures were used to calculate the MDSH composite score. Similar domain terms were grouped into one domain (i.e. “ Satisfaction” and “ Quality” were grouped into “ Satisfaction/Quality” domain, percentage of fast spindles, percentage of REM, spindle density (C3), and spindle density (C4) were grouped into “ Sleep Characteristics” domain, “ Alertness”, “ Sleepiness”, and “ Drowsiness” were grouped into “ Alertness/Sleepiness” domain, “ Timing” and “ Mid-sleep Timing” were grouped into “ Timing” domain, “ Continuity”, “ Sleep interruptions”, “ Wake after sleep onset”, “ Time awake”, and “ Awakenings” were grouped into “ Continuity” domain, and “ Insomnia”, Sleep disturbances, REM sleep behavior disorder”, “ Symptoms of sleep disorders”, “ Snoring”, and “ Average Oxygen Saturation” were grouped into “ Sleep Disorders” domain).

## 3. RESULTS

The search strategy generated 1,704 references, and 223 duplicates were removed. The 1,481 titles/abstract were reviewed, and 1,374 did not meet eligibility and were excluded. The 107 full-text articles were reviewed, and 68 were excluded due to not meeting eligibility. Thus, 39 eligible articles were included in the scoping review (Figure 1; Supplementary File 2). One study^15^ contained two MDSH composite scores; both are reported in this scoping review for a total of 40 MDSH composite scores.

**Figure 1.**
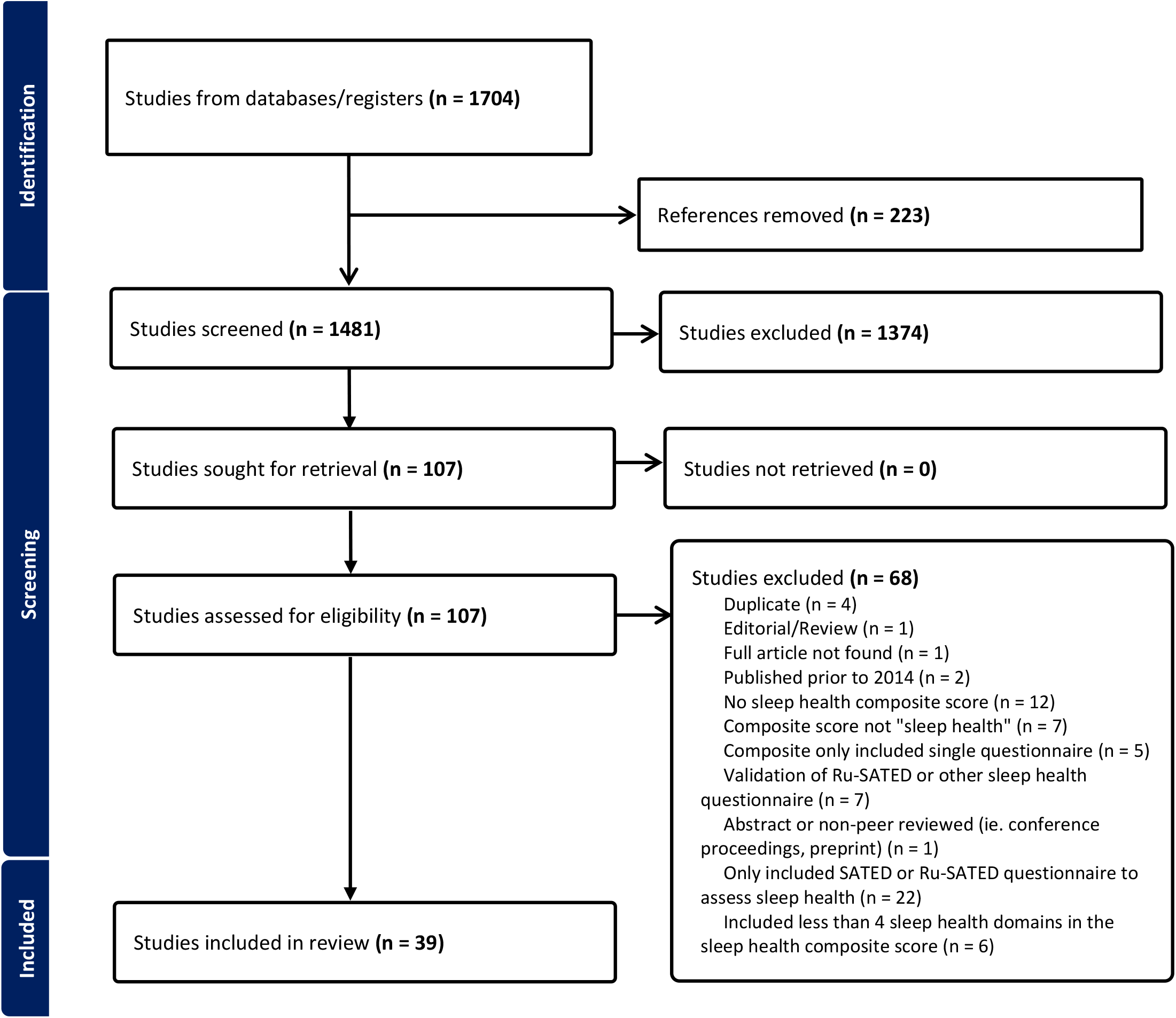
PRISMA diagram.

The majority of studies were conducted in the United States (n=31) followed by the United Kingdom (n=3), Japan and Australia (n=2 for both), and China (n=1; Table 1). Five and six domains were the most common number of domains included in the MDSH composite score (n=17 for both) followed by 7 domains (n=4), and two articles included 4 domains (Table 1). Seventeen articles included only self-report measures and n=19 articles included a mix of self-report and objective measures in their MDSH composite score (Table 1). Four articles included only objective measures in their MDSH composite score. One study did not specify the measurements used to assess the domains in their MDSH composite score.^16^

**Table 1.**
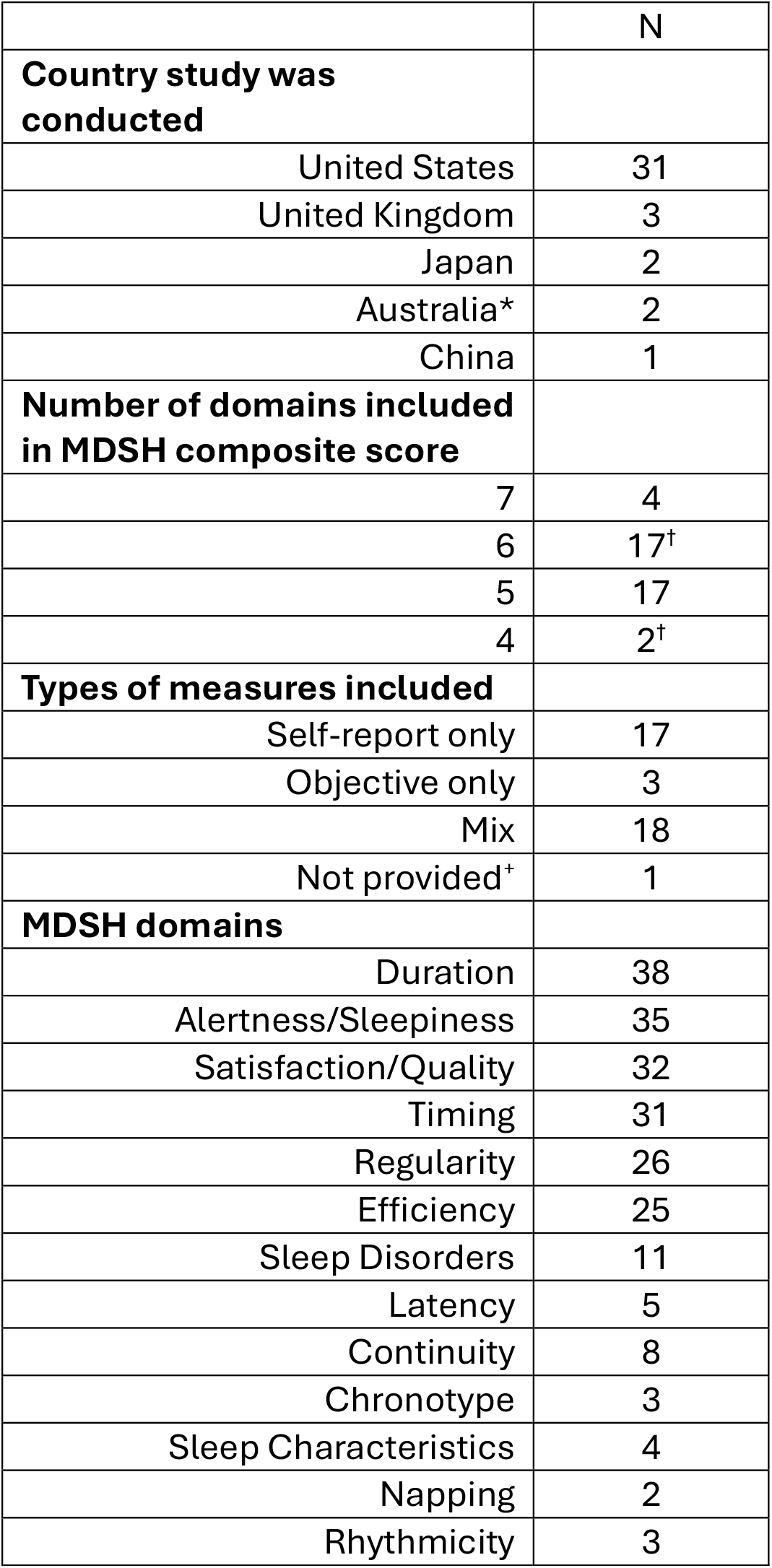
Countries studies conducted, number of domains included in the Multi-Dimensional Sleep Health (MDSH) score, type of measures included in the MDSH score, and the domains included in the MDSH scores. *Data included one article came from UK Biobank and authors in Australia. ^+^Original article did not specify the type of measure used to assess the domains. ^†^Yoo, et al. 2024 contains two MDSH composite scores; both are reported.

|  |  |
| --- | --- |
|  | N |
| **Country study was conducted** |  |
| United States | 31 |
| United Kingdom | 3 |
| Japan | 2 |
| Australia* | 2 |
| China | 1 |
| **Number of domains included in MDSH composite score** |  |
| 7 | 4 |
| 6 | 17<sup>†</sup> |
| 5 | 17 |
| 4 | 2<sup>†</sup> |
| **Types of measures included** |  |
| Self-report only | 17 |
| Objective only | 3 |
| Mix | 18 |
| Not provided<sup>†</sup> | 1 |
| **MDSH domains** |  |
| Duration | 38 |
| Alertness/Sleepiness | 35 |
| Satisfaction/Quality | 32 |
| Timing | 31 |
| Regularity | 26 |
| Efficiency | 25 |
| Sleep Disorders | 11 |
| Latency | 5 |
| Continuity | 8 |
| Chronotype | 3 |
| Sleep Characteristics | 4 |
| Napping | 2 |
| Rhythmicity | 3 |

Thirteen unique MDSH domains were identified (Table 1): Regularity, Satisfaction/Quality, Alertness/Sleepiness, Timing, Efficiency, Duration, Chronotype, Continuity, Latency, Napping, Rhythmicity, Sleep Disorders, and Sleep Characteristics. The domain most commonly included was Duration (n=38), followed by Alertness/Sleepiness (n=35), Satisfaction/Quality (n=32), and Timing (n=31). The domains were assessed in a variety of ways (Table 2; Supplementary File 3), with most domains (n=6) being assessed using both self-report and objective measures although some (n=4) were assessed using only self-report measures (i.e. Satisfaction/Quality) or only objective measures (i.e. Sleep Characteristics; n=3).

**Table 2.** Type of measure by MDSH domain. ^+^Article did not specify the type of measure used to assess the domains.

| **MDSH Domain** | **Type of Measures** |  |  |
| --- | --- | --- | --- |
|  | Self-report<br>(n) | Objective<br>(n) | *Not Reported<br>(n) |
| Duration | 17 | 20 | 1 |
| Alertness/Sleepiness | 34 | 0 | 1 |
| Satisfaction/Quality | 32 | 0 | 0 |
| Timing | 12 | 19 | 0 |
| Regularity | 7 | 19 | 0 |
| Efficiency | 8 | 17 | 0 |
| Sleep Disorders | 8 | 1 | 2 |
| Latency | 5 | 0 | 0 |
| Continuity | 4 | 4 | 0 |
| Chronotype | 2 | 0 | 1 |
| Sleep<br>Characteristics | 0 | 4 | 0 |
| Napping | 0 | 2 | 0 |
| Rhythmicity | 0 | 3 | 0 |

## 4. DISCUSSION

This scoping review describes the domains included in MDSH composite scores and how the domains are defined. Thirteen unique MDSH domains were identified, and how the domains were assessed varied. Articles tended to either use only self-report measures or a mix of self-report measures in their MDSH composite scores. In order to advance the study of sleep health, consensus is needed on the definition of sleep health domains or, at a minimum, clear reporting on the definitions used. Further research is needed to determine which MDSH domains are most associated with health outcomes, and thus most appropriate to include in a MDSH composite score.

It is perhaps not a surprise that the domains most commonly included were the domains within the Ru-SATED framework. The six most commonly included domains were Regularity (n=26), Satisfaction/Quality (n=32), Alertness/Sleepiness (n=35), Timing (n=31), Efficiency (n=25), and Duration (n=38). The SATED framework was proposed by Buysee^4^, and he provided evidence linking each sleep health domain when not achieved to negative health consequences. Buysse^4^ did discuss that Regularity was another dimension of sleep, and Ravyts et al.^10^ added Regularity as a sleep health domain, thus extending the framework to Ru-SATED in their study to evaluate the psychometric properties of the Ru-SATED scale.^10^ Thus, the SATED/Ru-SATED framework is a well-known sleep health framework. However, inclusion of seven other domains in MDSH composite scores included in this scoping review indicates a lack of consensus on what sleep health *is* and *how to measure* sleep health.

It is interesting that Sleep Disorders was included as a domain in MDSH composite scores 11 times considering sleep health is intended to be a positive attribute and *not* focused on sleep disorders or the absence of sleep disorders.^4^ Of the eleven times the Sleep Disorder domain was included, four instances included assessment of insomnia, three included assessment of sleep apnea, and one included assessment of sleep medications, REM Sleep Behavior Disorder, or symptoms of sleep disorders, and one did not provide details about the sleep disorder assessment. In four of these studies^16-19^, Sleep Disorder was included as a domain in the MDSH composite score two times. Perhaps including Sleep Disorders as a domain in the MDSH score is due to reluctance or resistance of the sleep field to move beyond the traditional medical model of focusing on sleep disorders.

There was a lot of variability in the particular measures used to assess various domains. For self-report measures, some articles used a single-item question while others used a validated sleep questionnaire. Articles that used objective assessments used actigraphy or polysomnography. It is reasonable that there are differences in methods used to assess the domains (ie. the Duration domain was assessed almost equally using self-report (n=17 instances) or objective measures (n=20 instances) whereas the Satisfaction/Quality domain was assessed only using self-report). However, it would be ideal to have consensus on the assessment (or range of assessments) for the respective domains. In addition, the direction of the MDSH composite score scales differed among studies (i.e. a higher number indicated better sleep health in some studies but indicated worse sleep health in other studies). Consensus on the direction of the scale is also needed to aid in interpretation and comparison between studies.

A limitation of this scoping review is articles were included if they defined the domains to be included in the MDSH composite a priori; we excluded articles that used modeling techniques to determine which sleep health domains should be included in a model or were predictive of a condition. For example, Buxton et al.^20^ included demographic variables, sleep apnea symptoms variables, and sleep health variables in regression models to predict cardiometabolic risk for workers in extended care and information technology industries. It may be that different domains should be included in a MDSH composite score depending on the health condition being targeted.

In conclusion, while the domains most often included in the MDSH composite scores followed the SATED/Ru-SATED framework, there was variability in the domains included as well as variability in how the domains were assessed. Further discussion is needed within the sleep health research and clinical community to reach consensus on what domains should be included in a MDSH composite score and the most appropriate assessment(s) to use to measure respective domains.

## Data Availability

All data produced in the present study are available upon reasonable request to the authors

## References

1. Research CoSMa. Sleep Disorders and Sleep Deprivation: An Unmet Public Health Problem. National Academies Press; 2006.

2. Di H, Guo Y, Daghlas I, et al. Evaluation of Sleep Habits and Disturbances Among US Adults, 2017–2020. JAMA Netw Open. Nov 1 2022;5(11):e2240788. doi:10.1001/jamanetworkopen.2022.40788

3. Foundation NS. Sleep in America Poll: Sleep Health & Scheduling. Accessed April 28, 2021, https://www.sleepfoundation.org/wp-content/uploads/2019/02/SIA_2019_Sleep_Health_and_Scheduling.pdf

4. Buysse DJ. Sleep health: can we define it? Does it matter? Sleep. Jan 1 2014;37(1):9–17. doi:10.5665/sleep.3298

5. Lobo A, Lopez-Anton R, de-la-Camara C, et al. Non-cognitive psychopathological symptoms associated with incident mild cognitive impairment and dementia, Alzheimer’s type. Neurotoxicity research. Oct 2008;14(2-3):263–72. doi:10.1007/BF03033815

6. Osorio RS, Pirraglia E, Aguera-Ortiz LF, et al. Greater risk of Alzheimer’s disease in older adults with insomnia. Journal of the American Geriatrics Society. Mar 2011;59(3):559–62. doi:10.1111/j.1532-5415.2010.03288.x

7. Hahn EA, Wang HX, Andel R, Fratiglioni L. A change in sleep pattern may predict Alzheimer disease. The American journal of geriatric psychiatry : official journal of the American Association for Geriatric Psychiatry. Nov 2014;22(11):1262–71. doi:10.1016/j.jagp.2013.04.015

8. Lim AS, Kowgier M, Yu L, Buchman AS, Bennett DA. Sleep Fragmentation and the Risk of Incident Alzheimer’s Disease and Cognitive Decline in Older Persons. Sleep. 2013;36(7):1027–1032. doi:10.5665/sleep.2802

9. Cricco M, Simonsick EM, Foley DJ. The impact of insomnia on cognitive functioning in older adults. Journal of the American Geriatrics Society. Sep 2001;49(9):1185–9.

10. Ravyts SG, Dzierzewski JM, Perez E, Donovan EK, Dautovich ND. Sleep Health as Measured by RU SATED: A Psychometric Evaluation. Behav Sleep Med. Dec 12 2019:1–9. doi:10.1080/15402002.2019.1701474

11. Wallace ML, Hall MH, Buysse DJ. Chapter 3 - Measuring sleep health. In: Nieto FJ, Petersen DJ, eds. Foundations of Sleep Health. Academic Press; 2022:37–71.

12. Tricco AC, Lillie E, Zarin W, et al. PRISMA Extension for Scoping Reviews (PRISMA-ScR): Checklist and Explanation. Ann Intern Med. Oct 2 2018;169(7):467–473. doi:10.7326/M18-0850

13. Peters MDJ, Marnie C, Tricco AC, et al. Updated methodological guidance for the conduct of scoping reviews. JBI Evid Implement. Mar 2021;19(1):3–10. doi:10.1097/xeb.0000000000000277

14. Barry A, Bock K, Vaduvathiriyan P, Siengsukon C. Domains included in multidimensional sleep health composites: A scoping review protocol. Accessed 3/4/25, https://osf.io/jpbmc

15. Yoo A, Vgontzas A, Chung J, et al. The association between multidimensional sleep health and migraine burden among patients with episodic migraine. Journal of clinical sleep medicine : JCSM : official publication of the American Academy of Sleep Medicine. Feb 1 2023;19(2):309–317. doi:10.5664/jcsm.10320

16. Huang BH, Hamer M, Duncan MJ, Cistulli PA, Stamatakis E. The bidirectional association between sleep and physical activity: A 6.9 years longitudinal analysis of 38,601 UK Biobank participants. Prev Med. Feb 2021;143:106315. doi:10.1016/j.ypmed.2020.106315

17. Sampasa-Kanyinga H, Chaput JP, Huang BH, Duncan MJ, Hamer M, Stamatakis E. Bidirectional associations of sleep and discretionary screen time in adults: Longitudinal analysis of the UK biobank. J Sleep Res. Apr 2023;32(2):e13727. doi:10.1111/jsr.13727

18. Schiel JE, Tamm S, Holub F, et al. Associations Between Sleep Health and Amygdala Reactivity to Negative Facial Expressions in the UK Biobank Cohort. Biological psychiatry. Nov 1 2022;92(9):693–700. doi:10.1016/j.biopsych.2022.05.023

19. Yu X, Deng S, Liu J, et al. Predictive Modeling Using a Composite Index of Sleep and Cognition in the Alzheimer’s Continuum: A Decade-Long Historical Cohort Study. J Alzheimers Dis Rep. 2024;8(1):589–600. doi:10.3233/ADR-240001

20. Buxton OM, Lee S, Marino M, Beverly C, Almeida DM, Berkman L. Sleep Health and Predicted Cardiometabolic Risk Scores in Employed Adults From Two Industries. Journal of clinical sleep medicine : JCSM : official publication of the American Academy of Sleep Medicine. Mar 15 2018;14(3):371–383. doi:10.5664/jcsm.6980

