## Supplementary File for "Domains included in multidimensional sleep health composite scores: A scoping review"

**Supplementary File 1: Complete Search Strategy**

| | |
|---|---|
| 1 | (multidimensional scor$ or multidimensional index or multidimensional measur$ or multidimensional scal$ or composit$ scal$ or composit$ scor$ or composit$ measur$ or composit$ index).ti,ab,kw. |
| 2 | ((sleep$ adj3 health) or (sleep$ adj3 qualit$)).ti,ab,kf. |
| 3 | 1 and 2 |
| 4 | (regularity or satisfaction or alertness or timing or efficiency or duration).ti,ab. and sleep.ti,ab,kw. and 1 |
| 5 | RU-SATED.ti,ab,kw. or (SATED and sleep$).ti,ab. |
| 6 | 4 or 5 |
| 7 | sleep/ or exp Sleep Wake Disorders/ or sleep deprivation/ or sleep duration/ or sleep quality/ or sleep latency/ or sleep stages/ |
| 8 | (sleep$ or sleep disorder$ or sleep deficien$ or sleep qualit$ or sleep behavior$ or sleep dysfunction$ or insomnia$ or sleepless$ or dyssomnia or parasomnia$ or somnolence).ti,ab,kw. |
| 9 | 7 or 8 |
| 10 | "Surveys and Questionnaires"/st or Self Report/st |
| 11 | 9 and 10 |
| 12 | (index or measure$ or survey$ or questionnaire$ or instrument$ or score$).ti,ab. and 9 and 1 |
| 13 | (monitoring, physiologic/ or actigraphy/ or actigraphy.ti,ab.) and 9 and 1 |
| 14 | Polysomnography/ or polysomnograph$.ti,ab. |
| 15 | actigraphy/ or actigraph$.ti,ab,kw. or wearables/ or wearable$.ti,ab,kw. |
| 16 | 14 and 15 |
| 17 | (is or mt or st).fs. |
| 18 | 16 and 17 |
| 19 | 3 or 6 or 11 or 12 or 13 or 18 |
| 20 | limit 19 to dt=20140101-20240531 |
| 21 | limit 20 to english language |

**Supplementary File 2: Full Reference List of Included Articles**


1. Bowman, M.A., et al., *Multidimensional sleep health is not cross-sectionally or longitudinally associated with adiposity in the Study of Women's Health Across the Nation (SWAN).* Sleep Health, 2020. **6**(6): p. 790-796.
2. Bowman, M.A., et al., *Longitudinal association between depressive symptoms and multidimensional sleep health: the SWAN sleep study.* Annals of Behavioral Medicine, 2021. **55**(7): p. 641-652.
3. Brindle, R.C., et al., *The relationship between childhood trauma and poor sleep health in adulthood.* Psychosomatic Medicine, 2018. **80**(2): p. 200-207.
4. Brindle, R.C., et al., *Empirical derivation of cutoff values for the sleep health metric and its relationship to cardiometabolic morbidity: results from the Midlife in the United States (MIDUS) study.* SLEEP, 2019. **42**(9).
5. Cavailles, C., et al., *Multidimensional Sleep Health and Long-Term Cognitive Decline in Community-Dwelling Older Men.* Journal of Alzheimer's Disease, 2023. **96**(1): p. 65-71.
6. Chen, T.Y., et al., *Poor sleep health predicts the onset of a fear of falling among community-dwelling older adults.* Sleep health, 2024. **10**(1): p. 137-143.
7. Chung, J., et al., *Multidimensional sleep health in a diverse, aging adult cohort: Concepts, advances, and implications for research and intervention.* Sleep health, 2021. **7**(6): p. 699-707.
8. Clementi, M.A., et al., *Preliminary exploration of a multidimensional sleep health composite in adolescent females with frequent migraine.* Headache, 2023. **63**(10): p. 1437-1447.
9. DeSantis, A.S., et al., *A preliminary study of a composite sleep health score: associations with psychological distress, body mass index, and physical functioning in a low-income African American community.* Sleep Health, 2019. **5**(5): p. 514-520.
10. Dong, L., et al., *Food insecurity, sleep, and cardiometabolic risks in urban American Indian/Alaska Native youth.* Sleep health, 2023. **9**(1): p. 4-10.
11. Dong, L., et al., *A composite measure of sleep health predicts concurrent mental and physical health outcomes in adolescents prone to eveningness.* Sleep health, 2019. **5**(2): p. 166-174.
12. Ensrud, K.E., et al., *Multidimensional sleep health and subsequent health-care costs and utilization in older women.* Sleep, 2020. **43**(2): p. zsz230.
13. Furihata, R., et al., *Association between a composite measure of sleep health and depressive symptoms in patients with obstructive sleep apnea treated with CPAP therapy: Real-world data.* Sleep Medicine, 2024. **120**: p. 22-28.
14. Furihata, R., et al., *An aggregate measure of sleep health is associated with prevalent and incident clinically significant depression symptoms among community-dwelling older women.* Sleep: Journal of Sleep and Sleep Disorders Research, 2017. **40**(3): p. 1-11.
15. Furihata, R., et al., *A composite measure of sleep health is associated with symptoms of depression among Japanese female hospital nurses.* Comprehensive Psychiatry, 2020. **97**.
16. Griggs, S., G. Pignatiello, and R.L. Hickman, *A composite measure of sleep health is associated with glycaemic target achievement in young adults with type 1 diabetes.* Journal of Sleep Research, 2023. **32**(3): p. 1-9.

17. Harvey, A.G., et al., *A randomized controlled trial of the Transdiagnostic Intervention for Sleep and Circadian Dysfunction (TranS-C) to improve serious mental illness outcomes in a community setting.* Journal of Consulting and Clinical Psychology, 2021. **89**(6): p. 537-550.
18. Hawkins, M.S., et al., *The association between multidimensional sleep health and gestational weight gain.* Paediatric and Perinatal Epidemiology, 2023. **37**(7): p. 586-595.
19. Hawkins, M.S., et al., *Associations between sleep health and obesity and weight change in adults: The Daily24 Multisite Cohort Study.* Sleep health, 2023. **9**(5): p. 767-773.
20. Huang, B.-H., et al., *The bidirectional association between sleep and physical activity: A 6.9 years longitudinal analysis of 38,601 UK Biobank participants.* Preventive Medicine, 2021. **143**: p. N.PAG-N.PAG.
21. Kline, C.E., et al., *The association between sleep health and weight change during a 12-month behavioral weight loss intervention.* International Journal of Obesity, 2021. **45**(3): p. 639-649.
22. Lee, S. and K.M. Lawson, *Beyond single sleep measures: A composite measure of sleep health and its associations with psychological and physical well-being in adulthood.* Social Science & Medicine, 2021. **274**.
23. Lee, S., et al., *Multidimensional Sleep Health Problems Across Middle and Older Adulthood Predict Early Mortality.* The journals of gerontology. Series A, Biological sciences and medical sciences, 2024. **79**(3).
24. Lee, S., et al., *Arthritis, Sleep Health, and Systemic Inflammation in Older Men.* Arthritis Care and Research, 2020. **72**(7): p. 965-973.
25. Madrid-Valero, J.J., et al., *Sleep in adults from the UK during the first few months of the coronavirus outbreak.* Journal of Sleep Research, 2022. **31**(2).
26. Makarem, N., et al., *Multidimensional sleep health is associated with cardiovascular disease prevalence and cardiometabolic health in US adults.* International journal of environmental research and public health, 2022. **19**(17): p. 10749.
27. Polanka, B.M., et al., *The association of multidimensional sleep health with adiposity in heart failure with preserved ejection fraction.* HEART & LUNG, 2023. **58**: p. 144-151.
28. Sampasa-Kanyinga, H., et al., *Bidirectional associations of sleep and discretionary screen time in adults: Longitudinal analysis of the UK biobank.* Journal of Sleep Research, 2023. **32**(2).
29. Savin, K.L., et al., *Social and built neighborhood environments and sleep health: The Hispanic Community Health Study/Study of Latinos Community and Surrounding Areas and Sueño ancillary studies.* Sleep: Journal of Sleep and Sleep Disorders Research, 2024. **47**(2): p. 1-12.
30. Schiel, J.E., et al., *Associations between sleep health and amygdala reactivity to negative facial expressions in the uk biobank cohort.* Biological Psychiatry, 2022.
31. Thomas, M.C., D.J. Buysse, and I. Soreca, *Same sleep disorder but different sleep patterns: individual differences in sleep health and depressive symptomatology in veterans with obstructive sleep apnea.* Sleep and Breathing, 2024. **28**(3): p. 1431-1435.
32. Tighe, C.A., et al., *Multidimensional sleep health and physical functioning in older adults.* Gerontology and Geriatric Medicine, 2021. **7**: p. 23337214211016222.
33. Turner, M., et al., *The multidimensional sleep health of individuals with multiple sclerosis and Huntington's disease and healthy controls.* Journal of Clinical Sleep Medicine, 2024. **20**(6): p. 967-972.

34. Wallace, M.L., et al., *Which Sleep Health Characteristics Predict All-Cause Mortality in Older Men? An Application of Flexible Multivariable Approaches.* Sleep, 2018. **41**(1): p. 01.
35. Whibley, D., et al., *A multidimensional approach to sleep health in multiple sclerosis.* Multiple Sclerosis and Related Disorders, 2021. **56**.
36. Woo, J., et al., *The Association of Multidimensional Sleep Health With HbA<sub>1c</sub> and Depressive Symptoms in African American Adults With Type 2 Diabetes.* PSYCHOSOMATIC MEDICINE, 2024. **86**(4): p. 307-314.
37. Xing, M., et al., *Development and validation of a novel sleep health score in the sleep heart health study.* European Journal of Internal Medicine, 2024. **09**: p. 09.
38. Yoo, A., et al., *The association between multidimensional sleep health and migraine burden among patients with episodic migraine.* Journal of Clinical Sleep Medicine, 2023. **19**(2): p. 309-317.
39. Yu, X., et al., *Predictive Modeling Using a Composite Index of Sleep and Cognition in the Alzheimer's Continuum: A Decade-Long Historical Cohort Study.* Journal of Alzheimer's Disease Reports, 2024. **8**(1): p. 589-600.

Supplementary File 3: Assessment details by domain

| | Self-Report or Objective | Specifically, how assessed (questionnaire/question/ device) | Possible Responses | Definition of good sleep health |
|---|---|---|---|---|
| **Satisfaction/Quality** | | | | |
| (N=32) | Self-report | "On most nights, how many hours do you sleep? How many hours of sleep do you need to feel rested?"<br>(N=1) | +Not provided | hours of sleep needed to feel rested did not exceeded average hours slept at night |
| | | "How would you rate the overall quality of your sleep in the last month?"<br>(N=1) | 1-very good 2-good 3-fair 4- poor 5-very poor | 1-very good<br>2-good |
| | | "How "satisfied" are you with your sleep?"<br>(N=1) | very good/good/usual/bad/very bad | very good, good, usual |
| | | "Do not get enough sleep"<br>(N=1) | Never (0); Rarely (1/mo); Sometimes (2-4/mo); Often (5-15/mo); Almost always (16-30/mo) | Never, Rarely, Sometimes |
| | | "Do you obtain enough rest during sleep?"<br>(N=1) | very sufficient/ sufficient/insufficient/ very insufficient/ uncertain | very sufficient, sufficient, uncertain |
| | | "How would you rate your sleep quality for the majority of nights during the past month?"<br>(N=1) | 1 = very good; 2 = fairly good; 3 = fairly bad; 4 = very bad | 1: very good |
| | | "Have trouble falling asleep./Wake up during the night and have difficulty going back to sleep./Wake up too early in the morning and be unable to get back to sleep./Feel unrested during the day, no matter how many hours of sleep you had."<br>(N=1) | +Not provided | 0 for all 4 items |
| | | Sleep Diary<br><br>(N=6) | "Overall quality of your sleep last night"<br>1 (very good) to 5 (very poor)<br><br>(N=2) | <2.8<br>Average sleep quality of ≥ 3.5 |
| | | | sleep satisfaction ranging from 1 "very bad" to 5 "very good", 3"neutral"<br>(N=1) | 4 "good" to 5 "very good" |
| | | | average diary-assessed "readiness," with scores ranging from 0 ("not at all") to 4 ("extremely") rested<br>(N=1) | "Moderately", "quite a bit", or "extremely" rested upon awakening |
| | | | (1=Very good, 5=Very poor)<br>(N=1) | Average <2.8 |
| | | | average "restedness upon awakening"- 0=not at all; 4 = extremely<br>(N=1) | "moderately," "quite a bit", or "extremely" rested upon awakening |
| | | Daily sleep log ratings | rated on an 11-point Likert scale (0="great", 10="terrible") | average sleep quality was rated as <5 ("good" to "great") |

| | | | | |
|---|---|---|---|---|
| | | (N=1) | | |
| | | Pittsburgh Sleep Diary<br><br>(N=1) | Average self-reported sleep quality from daily sleep diary, 0 (very bad)-100 (very good) scale | Median split: >66.66 |
| | | ISI<br>(N=1) | 0-28 | <10 |
| | | PSQI<br>PSQI: Question 6<br>PSQI: “How would you rate your sleep quality overall?"<br>Sleep Quality item<br>PSQI: “During the past month, how would you rate your sleep quality overall?”<br><br>(N=12) | very good, fairly good, fairly bad, very bad<br><br>0 (very good) to 3 (very bad)<br><br>(N=9) | "fairly good" or "very good"<br>(N=8) |
| | | | | ≤1<br>(N=1) |
| | | | Range 0-21<br>(N=1) | ≤ 5 |
| | | | PSQI Global Score<br>(N=1) | global score of ≤5 |
| | | | +Not provided<br>(N=1) | +Not provided |
| | | Women’s Health Initiative Insomnia Rating Scale<br><br>(N=1) | Likert subscale | "sound and restful" or "very sound or restful" |
| | | PROMIS Sleep Disturbance Scale<br><br>(N=2) | each item ranked using 5-point Likert scale (not at all to very much). Scores range from 8-40, with higher scores indicating greater severity of sleep disturbance. The raw scores on the 8 items are summed to obtain a total raw score, which is then transformed into a t-score | PROMIS < 56.36 |
| | | | The 8-item short version assesses sleep disturbance over the past 7 days, including restlessness, sleep quality, ability to fall and stay asleep, and refreshment following sleep using a 1–5 scale (not at all or never to very much or always). Scores range from 8 to 40, with higher scores indicating increased disturbance | +Not provided |

Supplementary File 3: Assessment details by domain

| | | | | |
|---|---|---|---|---|
| | | +Not provided<br>(N=1) | +Not provided | "getting enough hours of sleep to feel rested" |
| +Not provided indicates instances where the original article did not specify | | | | |
| **Latency** | | | | |
| **(N=7)** | Self-Report | “How long does it usually take you to fall asleep each night?”<br><br>“How long (in minutes) has it usually taken you to fall asleep each night?”<br><br>“How many minutes does it usually take you to fall asleep at bedtime?”<br><br>“In the last month, how long did it usually take to fall asleep each night?”<br><br>(N=4) | +Not provided<br><br>(N=1) | < 30 minutes<br><br>(N-4) |
| | | | Number of minutes it takes to fall asleep<br><br>(N=3) | |
| | | “Thinking about a typical night in the last month…How long does it take you to fall asleep?”<br><br>(N=1) | 1 = 0–15 min<br>2 = 16–30 min<br>3 = 31– 45 min<br>4 = 46–60 min<br>5 = >60 min | 1: 0-15min |
| | | Asked whether or not they ever told a doctor that they had trouble sleeping<br>(N=1) | Yes or No | responded that they have never told their doctor that they have trouble falling asleep |
| | | +Not provided<br>(N=1) | Number of minutes to fall asleep | taking <30 minutes to fall asleep |
| +Not provided indicates instances where the original article did not specify | | | | |
| **Timing** | | | | |
| **(N=30)** | Self-Report | Responses to wake up time and bedtime<br><br>(N=1) | Mid-sleep time was calculated using the following formula: "Mid-sleep time" = "bedtime" + ("wake up time" - "bedtime") / 2. Chronotypes were categorized based on quintile of the mid-sleep time, where early-chronotype was the 1st quintile (lowest score: b3:30 AM), intermediate-chronotype consisted of from 2nd, 3rd, and 4th quintile, and late-chronotype was the 5th quintile (highest score: ≥6:00)<br><br>(N=1) | 2nd, 3rd, 4th quintile |

Supplementary File 3: Assessment details by domain

| | | | | |
|---|---|---|---|---|
| | | At what time do you usually fall asleep? At what time do you usually wake up?<br><br>(N=2) | Assessed by identifying the clock time halfway between falling asleep time and waking up time and was calculated as the sum of falling asleep time + (waking up time - falling asleep time)/2. Mid-sleep time was categorized based on octiles of the mid-sleep time.<br><br>(N=1) | Women in the 2nd thru 7th octile |
| | | | Clock time<br><br>(N=1) | 2-3:59am |
| | | "In the last month, what time did you usually go to bed at night?"<br>"In the last month, what time did you usually get up in the morning?"<br><br>(N=1) | Mid-sleep time (ie, middle point between the time when individuals went to bed and the time when individuals got up): Hour/Minute/AM/PM<br><br>(N=1) | Between 2 am to 3:59am |
| | | "What time have you usually gone to bed at night? How long (in minutes) has it usually taken you to fall asleep each night? What time have you usually gotten up in the morning?"<br><br>(N=1) | Sleep onset time was calculated using the following formula: "sleep onset time" = "bedtime" + "sleep onset latency." Mid-sleep time was calculated based on responses to wake time<br>and sleep onset time using the following formula: "mid- sleep time" = "sleep onset time" + ("wake time" - "sleep onset time")/ 2<br><br>(N=1) | 2-4am<br><br>(N=1) |
| | | Sleep Diary<br><br>(N=3) | average mid-point of sleep<br>(N=2) | 2-4am<br>(N=2) |
| | | | Mean midpoint<br>(N=1) | +Not provided<br>(N=1) |
| | | Pittsburgh Sleep Diary<br><br>(N=1) | Average of calculated sleep midpoint from daily sleep diary, midnight = 0 min<br>(N=1) | Male: ≥147 OR ≤ 218 min<br>Female: ≥161 OR ≤ 234 min |
| | | Morningness-Eveningness Questionnaire<br><br>(N=1) | The scores on the 19 items are summed to obtain a total score. Scores range from 16-86 | 42-58 |

Supplementary File 3: Assessment details by domain

| | | | | |
|---|---|---|---|---|
| | | | (N=1) | |
| | | MCTQ<br>(N=1) | midpoint between sleep onset and sleep offset.<br>(N=1) | <4:00am |
| | | +Not provided<br><br>(N=1) | Mid-sleep time, calculated as the midpoint of the in-bed interval. Mid-sleep time was categorized based on octiles<br><br>(N=1) | The second-seventh octiles combined |
| | Objective | Actigraphy<br>(N=19) | Midpoint was the clock time midway between sleep onset and offset. Sleep midpoint was converted to minutes before/after 4:00 AM (approximate median midpoint of the sample) to allow for linear analysis.<br>(N=1) | 2:00 am - 4:00 am |
| | | | Sleep midpoint represented by the middle of the sleep period between the sleep onset and final awakening<br><br>midpoint from sleep onset to wake up (hh:mm)<br><br>mean sleep midpoint (midpoint of bed and wake time)<br><br>average mean sleep midpoint<br><br>mean actigraphic sleep midpoint<br><br>the average midpoint of sleep, calculated as the difference between sleep onset and wake time divided by two, and adding this value to the time of sleep onset (sleep onset + [{wake time sleep onset}]<br><br>sleep midpoint, occurring midway between sleep onset and offset | 2:01am-4:00am<br><br>Sleep midpoint of 2:01-04:00AM<br>(N=4) |
| | | | | Before 4am<br>(N=2) |
| | | | | 2:00am-4:00am<br><br>Average sleep midpoint between 2:00am and 4:00am<br><br>(N=8) |
| | | | | average sleep midpoint before 4 am<br>(N=1) |
| | | | | 2:24am-3:30am<br><br>(N=3) |

Supplementary File 3: Assessment details by domain

| | | | | |
|---|---|---|---|---|
| | | | Sleep midpoint<br><br>average midpoint sleep<br><br>(N=18) | |
| *** 1 article has multiple instances (Yoo et al)** +Not provided indicates instances where the original article did not specify; MCTQ = Munich Chronotype Questionnaire | | | | |
| **Alertness/ Sleepiness** | | | | |
| **Alertness**<br><br>**(N=21)** | Self-Report | "In the last month, how often did you have trouble staying awake at times during the day when you wanted to be awake?"<br><br>(N=1) | 1-everyday (7 days a week)<br>2-most days (5-6 days a week)<br>3-some days (2-4 days a week)<br>4-rarely (once a week or less)<br>5-never | 4-rarely (once a week or less) and 5-never |
| | | During a usual week, how many times do you nap for 5 min or more?<br>(N=1) | +Not provided | ≤ 2 naps/wk |
| | | Typical Nap duration<br><br>(N=1) | Participants who reported napping at least once per week were asked how long their napes typically lasted: 1-30 minutes, 30-60 minutes, 60-90 minutes, or greater than 90 minutes. | <60 minutes min/day |
| | | Sleep diary; "Did you nap today?"<br><br>(N=1) | Yes or no: Summed yes responses across the week and dichotomized to: no alertness (≥ nap frequency; coded as 1) or alertness (<3 nap frequency; coded as 0). | <3 nap frequency |
| | | Sleep Diary; "How alert were you today?"<br>(N=2) | 1=Most alert, 5=Not at all alert | Average <2.2 |
| | | Epworth Sleepiness Scale (ESS)<br><br>(N=9) | Total score 0-24<br>Range 0-24<br>(N=7) | 13 or less<br>(N=1) |
| | | | | total score ≤10<br>≤10<br>(N=6) |
| | | | Scores range from 0-24, with higher scores indicating higher sleepiness<br>(N=1) | ≤ 7.5 |
| | | | +Not provided<br>(N=1) | ≤10 |
| | | PSQI: "How often have you had trouble staying awake while driving, eating meals, or engaging in social activity?"<br>(N=1) | 0 (not during the past month) to 3 (3 or more times a week) | 0=not during the past month |

Supplementary File 3: Assessment details by domain

| | | | | |
|---|---|---|---|---|
| | | AIQ item "I feel sleepy or tired during the day"<br>(N=1) | 5-point Likert scale ranging from 0 (“never”) to 4 (“almost always”) | rated this item as ≤2 (“never” to “sometimes”) |
| | | 2 items in the School Sleep Habits Survey for Adolescents (fallen asleep in a morning class in the past 2 weeks, and fallen asleep in an afternoon class in the past 2 weeks)<br>(N=1) | 0 (never), 1 (once), 2 (twice), 3 (several times), to 4 (every day/night) | responded "never" to both questions. |
| | | Sleepiness Scale<br>(N=1) | total score on the 10-item Sleepiness Scale | ≤7.5 |
| | | Promis SRI T-score<br>(N=1) | 8-item shortform version | PROMIS SRI T-score ≤ 50 |
| | | Day-time sleepiness question on PROMIS-SRI<br>(N=1) | 16-item version assessed with a 5-point scale | +Not provided |
| **Sleepiness (N=6)** | Self-Report | “How likely are you to doze off or fall asleep during the daytime when you don't mean to? (e.g. when working, reading, or driving)”<br>(N=1) | "never/rarely" "sometimes" "often" "all of the time" | "never/rarely" |
| | | Feels excessively (overly) sleepy during the day<br>(N=1) | Never (0); Rarely (1/mo); Sometimes (2-4/mo); Often (5-15/mo); Almost always (16-30/mo) | Never, Rarely, Sometimes |
| | | ESS<br>(N=2) | range 0-24<br>Total Score | ≤10<br>(N=2) |
| | | +Not provided+<br>(N=2) | +Not provided | Infrequent<br>infrequent daytime sleepiness |
| **Drowsiness (N=1)** | Self-Report | ESS<br>(N=1) | +Not provided | +Not provided |
| **Combination*<br><br>(N=7)** | Self-Report | “How often they felt excessive daytime sleepiness”<br><br>(N=1) | never, rarely (once/month)<br>sometimes (2-4 times/month)<br>often (5-15 times/month)<br>almost always (16-30 times/month) | reported never or rarely having excessive daytime sleepiness |
| | | “Do you feel excessively sleep during the daytime?”<br>(N=1) | never/seldom/ sometimes/often/always | +Not provided |
| | | ESS<br><br>(N=5) | 0-24<br>(N=4)<br>+Not provided<br>(N=1) | ≤10<br><br>(N=5) |
| *Combination refers to domains that were referred to in the original article as both alertness and sleepiness; +Not provided indicates instances where the original article did not specify further; ESS = Epworth Sleepiness Scale | | | | |
| **Sleep Characteristics** | | | | |
| (N=1) | Objective | PSG<br>Percentage of fast spindles (%) | <30 (0)<br>30-50 (1) | <30 and 50-65 |

| | | | | |
|---|---|---|---|---|
| | | | 50-65 (0)<br>≥65 (2) | |
| | | PSG<br>Percentage of REM (%) | <15 (3)<br>15-20 (1)<br>20-25 (2)<br>≥25 (0) | ≥25 |
| | | PSG<br>Spindle Density (C3) | 0-0.5 (2)<br>0.5-1.0 (2)<br>1.0-1.5 (2)<br>≥1.5 (0) | ≥1.5 |
| | | PSG<br>Spindle Density (C4) | 0-0.5 (4)<br>0.5-1.0 (1)<br>1.0-1.5 (0)<br>≥1.5 (0) | 1.0-1.5 and ≥1.5 |
| PSG=polysomnography | | | | |
| **Continuity** | | | | |
| **Awakenings**<br><br>**(N=1)** | Self-Report | "How many times did you wake up between the time you first fell asleep and your final awakening"<br><br>(N=1) | 0 = 0 awakenings; 1 = 1 awakening, 2 = 2 awakenings,<br>3 = 3 awakenings, 4 = 4 or more awakenings | 0 awakenings |
| **Time awake**<br><br>**(N=1)** | Self-Report | "If you then wake up during the night, how long are you awake in total (add up all the time when you are awake)?"<br><br>(N=1) | 1 = 0–15 min, 2 = 16–30 min; 3 = 31–45 min, 4 = 46–60 min, 5 = >60 min | 0-15min |
| **Continuity**<br><br>**(N=4)** | Self-report | Sleep interruptions: MCTQ<br>(N=1) | frequency of sleep interruptions per week | no sleep interruptions |
| | | Sleep Onset Latency: MTCQ<br>(N=1) | Number of minutes to fall asleep | ≤ 30 min |
| | Objective | Actigraphy<br><br>(N=3) | Average wake-after-sleep-onset | < 40.93 |
| | | | WASO; the number of minutes an individual is awake after falling asleep | < 88 |
| | | | Actigraphy-based mean time awake after sleep onset (min) | < 44.94 |
| **Wake after sleep onset**<br><br>**(N=1)** | Objective | Actigraphy<br><br>(N=1) | total amount of wakes in bed (after sleep onset) per night (in minutes) and calculated average | <90 minutes/night |

Supplementary File 3: Assessment details by domain

| | | | | |
|---|---|---|---|---|
| *1 article has multiple instances (Hawkins 2023); MCTQ =Munich Chronotype Questionnaire | | | | |
| **Napping/Nap/Nap duration** | | | | |
| **(N=2)** | Objective | Actigraphy<br><br>(N=2) | weekly frequency; Nap was defined as out-of-bed sleep for 5 consecutive minutes or more | <2 times/week |
| | | | the average duration of naps | <100min/day |
| **Duration/ Nighttime sleep duration** | | | | |
| **(N=37)** | Self-report | "In the last month, how many hours of actual sleep did you usually get at night? This may be different than the number of hours you spent in bed."<br><br>"How many hours of actual sleep did you get at night?"<br><br>"How many hours of sleep do you usually get at night?"<br><br>"On average, how many hours do you sleep?"<br><br>"On most nights, how many hours do you sleep?"<br><br>"How much sleep do you usually get at night (or in your main sleep period) on weekdays or workdays?"<br><br>(N=6) | Number of hours<br><br>(N=6) | ≥7 hours to < 9 hours<br>(N=3) |
| | | | | ≥6 and ≤8 h<br>(N=1) |
| | | | | 6-8 h<br>(N=1) |
| | | | | >6 hours<br><br>(N=1) |
| | | Reported the number of hours they usually sleep on weekdays/workdays and on weekends/non-workdays.<br><br>(N=1) | The weekday-weekend difference in sleep duration was computed | ≥ 7h and < 9 h |
| | | About how many hours sleep do you get in every 24 hours? (please include naps).<br>(N=1) | short sleepers (<7 hours),<br>normal sleepers (7-9 hours),<br>long sleepers (>9 hours) | 7-9 hours |

Supplementary File 3: Assessment details by domain

| | | | | |
|---|---|---|---|---|
| | | PSQI<br><br>(N=2) | "During the past month, how many hours of actual sleep did you get at night? This may be different than the number of hours you spend in bed."<br>(N=1) | 7-9 hr |
| | | | "During the majority of the days and nights in the past month how many hours of actual sleep did you get at night? This may be different to the number of hours you spent in bed" (1 = 7 to >9 hr; 2 = 6–7 hr; 3 = 5–6 hr; 4 = <5 hr)<br>(N=1) | 1 = 7 to >9 hr |
| | | Sleep Diary<br>(N=3) | Total sleep time; Sleep duration<br>(N=1) | +Not provided |
| | | | Total sleep time average<br>(N=1) | 9-11 h for 10-13 y and 8-10 h for 14-18 y |
| | | | Average duration | between 420 and 540 minutes (7–9 hours) |
| | | Difference in sleep onset and offset on the MCTQ<br><br>(N=1) | Number of Hours | 7-9 hr/night |
| | | Pittsburgh Sleep Diary<br><br>(N=1) | Average self-reported sleep duration from daily sleep diary | 7-8 hr<br><br>(N=2) |
| | | +Not provided<br><br>(N=1) | Number of hours | |
| | | +Not provided<br><br>(N=1) | +Not provided | 7-8 h/day |
| | | +Not provided<br><br>(N=1) | +Not provided | Adequate sleep duration (7-8 h/d) |
| | Objective | Actigraphy<br><br>(N=20) | Total time in minutes spent asleep between sleep onset and offset during the main sleep period<br><br>(N=1) | ≥6 hours |

Supplementary File 3: Assessment details by domain

| | | | | |
|---|---|---|---|---|
| | | | Minutes per night<br>(N=1) | >6 hours/night |
| | | | Defined as the total amount of minutes scored as main sleep in a given day<br>(N=1) | 6 hr < or = duration < or = 8 hr |
| | | | Average duration<br>(N=1) | between 420 and 540 minutes (7–9 hours) |
| | | | Number of minutes<br><br>total sleep time in minutes<br><br>minutes of actual asleep at night<br><br>mean total sleep time (min)<br><br>(N=6) | 320 and 426 total minutes between sleep onset and wake onset excluding wakefulness<br>(N=1) |
| | | | | 320minutes to 426minutes<br>(N=1) |
| | | | | >358.96 and <451.17<br>(N=1) |
| | | | | total sleep time ≥420 minutes (7 hours)<br>(N=1) |
| | | | | 319.6-450.3<br>(N=1) |
| | | | | 419.57-515.5<br>(N=1) |
| | | | Number of hours<br>Total amount of sleep obtained (hh:mm)<br>Average total sleep time<br>Average total minutes of sleep<br>Total sleep time<br>Mean actigraphic total sleep time<br>Total sleep time average<br>Total sleep time during the main sleep period<br>Average actigraphy calculated sleep duration<br><br>(N= 10) | 6-8 hours<br>(N=6) |
| | | | | 5h 20min-7h 6min<br>(N=1) |
| | | | | Participants aged 12–13 years: 9–11h. Participants aged 14–18years: 8–10h.<br>(N=1) |
| | | | | 7-9 h/night<br>(N=1) |
| | | | | Between 8-10 hours<br>(N=1) |
| *** 1 article has multiple instances (Yoo et al);** MCTQ = Munich Chronotype Questionnaire | | | | |
| **Efficiency** | | | | |

Supplementary File 3: Assessment details by domain

| | | | | |
|---|---|---|---|---|
| **(N=22)** | Self-report | "How long does it usually take you to fall asleep at bedtime?"<br>(N=1) | +Not provided | ≤30 min |
| | | Sleep diary<br>(N=3) | average sleep efficiency<br>(N=2) | ≥85%<br>(N=1) |
| | | | | ≥ 90.8<br>(N=1) |
| | | | Mean and variability calculated as total sleep time/time in bed ×100<br>(N=1) | ≥85%<br>(N=1) |
| | | PSQI<br>(N=1) | Derived from 3 items on PSQI: sleep duration, getting up time and bedtime; calculated (sleep duration/time in bed) X 100 | ≥85% |
| | | Pittsburgh Sleep Diary<br>(N=1) | Average calculated sleep efficiency | >85% |
| | Objective | Actigraphy<br><br>(N=16) | The amount of time in minutes awake between sleep onset and offset in the main sleep period (WASO)<br>(N=1) | <60 minutes |
| | | | Average sleep efficiency<br>(N=1) | ≥ 88.6 |
| | | | average sleep efficiency<br><br>mean actigraphic sleep efficiency<br><br>(N=3) | ≥85%<br><br>(N=3) |
| | | | ratio of total sleep time to time in bed (%)<br><br>total sleep time divided by total time in bed<br><br>total sleep time divided by total time in bed multiplied x100<br><br>total duration of objectively measured sleep divided by the total time in bed<br><br>(N= 4) | ≥85%<br><br>(N=4) |

| | | | | |
|---|---|---|---|---|
| | | | % total time in bed asleep<br>(N=1) | >85% |
| | | | calculated as the ratio of time spent asleep to total time spent in bed, multiplied by 100 (expressed as a percentage)<br>(N=1) | ≥85% percentile |
| | | | the average total minutes of sleep following sleep onset (first 1-minute epoch scored as sleep) divided by time in bed, multiplied by 100)<br>(N=1) | Greater than 85% |
| | | | Total time spent asleep divided by total time in bed, multiplied by 100<br><br>Total sleep time divided by total sleep period<br><br>(N=2) | >83%<br><br>(N=2) |
| | | | Average actigraphy calculated sleep efficiency<br>(N=1) | >83.0% |
| | | | Sleep maintenance efficiency<br>(N=1) | >90% |
| | | PSG<br>(N=1) | ≥80 (0)<br><80 (3)<br>(N=1) | ≥80 |
| *** 1 article has multiple instances (Yoo et al)** +Not provided indicates instances where the original article did not specify; PSQI = Pittsburg Sleep Quality Index; PSG=Polysomnography | | | | |
| **SLEEP DISORDERS** | | | | |
| **Insomnia**<br><br>**(N=4)** | Self-report | "Do you have difficulty falling asleep?": difficulty initiating sleep (DIS)<br>"Do you wake up frequently at night?": difficulty maintaining sleep (DMS)<br>"Do you wake up too early in the morning?": early morning awakening (EMA)<br><br>(N=1) | never/seldom/sometimes/often/always | Answering never, seldom, or sometimes to all three questions |

Supplementary File 3: Assessment details by domain

| | | | | |
|---|---|---|---|---|
| | | Do you have trouble falling asleep at night or do you wake up in the middle of the night?<br><br>(N=1) | "never/rarely" "sometimes" "usually" | "never/rarely" and "sometimes" |
| | | +Not provided<br>(N=2) | +Not provided | never or rare insomnia |
| **Sleep apnea**<br><br>**(N=3)** | Self-report | +Not provided<br>(N=2) | +Not provided | never or rare snoring |
| | Objective | PSG<br>Average Oxygen Saturation<br>(N=1) | <90 (8)<br>90-95 (2)<br>95-100 (0) | 95-100% |
| **REM sleep behavior disorder (RBD)**<br>**(N=1)** | Self-report | Participants reported the frequency with which they snore and snort (or stop breathing during their sleep)<br><br>(N=1) | never, rarely (1-2 nights/wk), occasionally (3-4 nights/wk), and frequently (5+ nights/wk) | never or rarely snoring, snorting, or having difficulty breathing during sleep |
| **Sleep disturbances**<br>**(N=1)** | Self-report | +Not provided<br>(N=1) | +Not provided | +Not provided |
| **Symptoms of sleep disorders**<br>**(N=1)** | Self-report | RBDSQ<br><br>(N=1) | +Not provided | +Not provided |
| **Sleep medication use**<br>**(N=1)** | Self-report | List given to participants (sedatives and hypnotics) and self-reported to a nurse to assess sleep medication use.<br><br>(N=1) | Yes or No | No |
| ***4 articles have two instances in the domain (Sampasa-Kanyinga 2023, Schiel 2022, Yu 2024, Huang 2021)** +Not provided indicates instances where the original article did not specify; PSG = Polysomnography; RBDSQ = REM Sleep Behavior Disorder Screening Questionnaire | | | | |
| **Regularity** | | | | |
| **(N=25)** | Self-report | Reported the times they usually fall asleep and wake up on weekdays/workdays and on weekends/non-workdays<br>(N=1) | The weekday-weekend difference in sleep midpoint (i.e., sleep timing) was computed | <2 h difference |
| | | Sleep Diary<br><br>(N=3) | midpoint sleep fluctuation (standard deviation for midpoint) | ≤1 h |
| | | | Sleep Regularity Index | ≥ 76.3 |
| | | | midpoint fluctuations | +Not provided |

Supplementary File 3: Assessment details by domain

| | | | | |
|---|---|---|---|---|
| | | Pittsburgh Sleep Diary<br>(N=1) | Standard deviation of calculated sleep midpoint | Male: ≤29 min Female: ≤26 min |
| | | +Not provided<br>(N=2) | Difference in sleep midpoint between work and work-free days after adjusting for potential make-up sleep on non-workdays (ie. social jetlag) | <60 minutes |
| | | | Difference between workday sleep duration and nonworkday sleep duration | Absolute value ≤60 min |
| | Objective | Actigraphy<br><br>(N=19) | Sleep Regularity Index<br>(N=1) | ≥ 68.0 |
| | | | Interdaily stability; regularity of sleep-wake patterns. Calculated as proportion of the total variance in sleep-wake status (dichotomized) at a particular epoch explained by clock time. Can range from 0- (the clock time explains none of the variance) to 1 (the clock time explains all of the variance.)<br>(N=1) | ≥0.739 |
| | | | Standard deviation in wake time values<br>(N=1) | <0.84<br>(N=1) |
| | | | Standard deviation of wake time Standard deviation of wake time (hrs)<br>(N=2) | < 0.75<br>(N=1) |
| | | | | <1.17<br>(N=1) |
| | | | within-person standard deviation in nightly sleep duration<br>(N=1) | nightly sleep duration SD was ≤1h |
| | | | standard deviation of sleep duration<br>(N=1) | < 60 min |
| | | | standard deviation of actigraphic waketime<br>(N=1) | < 60 min |
| | | | Standard deviation of calculated sleep midpoint | < 60 min<br>(N=2) |
| | | | | ≤ 1 h |

| | | | | |
|---|---|---|---|---|
| | | | SD of midpoint of sleep (hh:mm)<br><br>standard deviation of the individual's sleep midpoint<br><br>standard deviation from sleep midpoint<br><br>Sleep midpoint standard deviation<br><br>midpoint standard deviation<br>(N=6) | (N=1)<br><br>< 65 minutes<br>(N=3) |
| | | | The variability in sleep midpoint across the week, quantified using standard deviation<br>(N=1) | <1 h 5 min<br>(N=1) |
| | | | midpoint variability (SD, min)<br>(N=1) | < 30 min<br>(N=1) |
| | | | SD of sleep midpoint<br><br>average within-person standard deviation<br><br>(N=3) | ≤ 1 SD<br>(N=1)<br><br><60 min<br><br>(N=2) |
| *** 1 article has multiple instances (Yoo et al)** +Not provided indicates instances where the original article did not specify | | | | |
| **Chronotype** | | | | |
| **(N=3)** | Self-report | Do you consider yourself to be definitely a 'morning' person/more a 'morning' than an 'evening' person/more an 'evening' than a 'morning' person/definitely an 'evening' person?<br><br>(N=1) | The 2 middle responses were collapsed into an intermediate chronotype category, permitting comparisons with the early ("definitely morning") and late ("definitely evening") chronotype category. (also called early, intermediate, late) | Early |
| | | +Not provided<br><br>(N=2) | +Not provided<br><br>(N=2) | Morning chronotype<br><br>(N=2) |
| +Not provided indicates instances where the original article did not specify | | | | |

Supplementary File 3: Assessment details by domain

| **Rhythmicity** | | | | |
|---|---|---|---|---|
| **(N=3)** | Objective | Actigraphy<br><br>(N=3) | The probability of being asleep or awake at the same time 24 hours apart | > 52.59 |
| | | | Pseudo-F statistics (PsF) | > 785.60 |
| | | | Probability of being in same state at any 2 time points 24 h apart | <0.69 |

Supplementary File 4: Data Extraction

| Last name of first author | Publication Year | Country Study Conducted | How was the sleep health composite calculated/ Scale description | How were the sleep health domains assessed | Sample Size | Age | Ethnicity Race | Sex | Number of Domains | Domains Included |
|---|---|---|---|---|---|---|---|---|---|---|
| Bowman | 2020 | United States | Each continuous sleep health variable was dichotomized using the cut-points detailed in with 0 indicating poor and 1 indicating good sleep health, and these variables were summed to create a composite score ranging from 0 (poor) to 6 (good).<br><br>The scale ranges from 0-6 with higher score indicating good sleep health | mix of self-report and objective | 221 | 52.2 (2.1) | White n=110 (49.8%), African American n=67 (30.3%), Chinese n=44 (19.9%)<br><br>("race/ethnicity" from Table 2) | 100% Female | 6 | Satisfaction/Quality<br>Timing<br>Alertness/Sleepiness<br>Duration<br>Efficiency<br>Regularity |
| Bowman | 2021 | United States | Each sleep health variable was dichotomized using an empirical cutoff, with 0 indicating poor sleep health and 1 indicating good sleep health.<br><br>The six sleep health components were summed to create a sleep health score ranging from 0 to 6, with higher values indicating better sleep health | mix of self-report and objective | 302 | Age at SWAN baseline: 46.2 (2.7); Age at SWAN Sleep Study: 52.1 (2.1) | "Race/ethnicity" (Table 2) White 140 (46.2), African American 112 (37.1), Chinese American 50 (16.6) | 100% Female | 6 | Satisfaction/Quality<br>Timing<br>Alertness/Sleepiness<br>Duration<br>Efficiency<br>Regularity |
| Brindle | 2018 | United States | Each component of sleep health was dichotomized as 0 or 1, with a higher score indicating better sleep health.<br><br>Total sleep health scores range from 0 to 6, with a value of 6 indicating optimal sleep health and a value of 0 indicating poorest sleep health | self-report only | 161 | 59.85 (9.06) | 98.1% non-Hispanic white | 67.7% female | 6 | Satisfaction/Quality<br>Timing<br>Alertness/Sleepiness<br>Duration<br>Efficiency<br>Regularity |
| Brindle | 2019 | United States | The larger MIDUS II sample was used to derive cutoff values. In the MIDUS Refresher sample, sleep health scores were calculated using cutoff values derived from the MIDUS II sample. All sleep health | mix of self-report and objective | MIDUS II n=432; MIDUS refresher n=268 | MIDUS II 56.95 mean (SD 11.5), range 35-85; MIDUS Refresher 51.68 mean (SD 12.7), range 26-77 | MIDUS II: White n=299 (69.2%), Black n=119 (27.5%), Other n=14 (3.2%); MIDUS refresher: | MIDUS II: male n=172 (39.8%; MIDUS refresher: male n=117 (43.7%) | 6 | Satisfaction/Quality<br>Timing<br>Alertness/Sleepiness<br>Duration<br>Efficiency<br>Regularity |

| | | | | | | | | | | |
|---|---|---|---|---|---|---|---|---|---|---|
| | | | dimensions were first centered using the respective cutoff value, producing a new variable that could be interpreted as “distance” from the cutoff value. Next, all sleep health dimensions were transformed to z-scores to achieve a uniform scale across the dimensions. Z-scores were coded such that greater values indicated better sleep health. Finally, all six of the individual sleep health dimensions z-scores were summed, creating a total sleep health score.<br><br>An increasing positive total sleep health score indicates better sleep health, and an increasing negative score, poor sleep health | | | | White n=175 (65.3%), Black n=72 (26.9%), other n=21 (7.8%) | | | |
| Cavailles | 2023 | United States | Summing the number of "poor" dimensions.<br><br>Total scores ranging from 0 to 5 and higher scores indicating poorer sleep health. The scores were categorized according to the number of poor sleep health dimensions into two categories (1 versus 0) and three categories (0, 1-2, 3-5). | self-report only | 2811 | 76.0 ± 5.3 | not reported (race broken down in table 1 after references, but not for the entire sample) | 100% men | 5 | Satisfaction/Quality<br>Efficiency<br>Timing<br>Alertness/Sleepiness<br>Duration |
| Chen | 2024 | United States | A sleep health composite score was calculated by adding up the "poor" sleep health dimensions<br><br>0-5; higher score indicating poorer sleep health | Self-report only | 686 | 65-74, 55%<br>75-84, 36%<br>85+, 9% | White, 85%<br>Black, 6%<br>Other, 9% | Female, 53%<br>Male, 47% | 5 | Satisfaction/Quality<br>Latency<br>Timing<br>Alertness/Sleepiness<br>Duration |
| Chung | 2021 | United States | Cut-points defined optimal ranges, with coding as "favorable" sleep as "1" and nonoptimal ranges as "0"<br><br>The scale ranges from 0-6 with a higher score indicating better sleep health | mix of self-report and objective | 735 | 59.4 (3.0) | Race-ethnicity (Table 2): White n=265 (36.1%); Chinese n=93 (12.7%); Black n=200 (27.2%); Hispanic n=177 (24.1%) | female n=409 (55.6%) | 6 | Satisfaction/Quality<br>Timing<br>Alertness/Sleepiness<br>Duration<br>Efficiency<br>Regularity |
| Clementi | 2023 | United States | Sum of all domains | mix of self- | 55 | 16.03 (1.64) | Hispanic/Latin o n=18 (33%) | 100% Female | 6 | Satisfaction/Quality<br>Timing |

| | | | | | | | | | | |
|---|---|---|---|---|---|---|---|---|---|---|
| | | | Scale ranging from 0-6 with higher scores indicating poorer sleep health | report and objective | | | Non-Hispanic/ Latino n=37 (67%)<br>Black or African American n=2 (4%)<br>More than one race n=15 (27%)<br>White n=38 (69%) | | | Alertness/Sleepiness<br>Duration<br>Efficiency<br>Regularity |
| DeSantis | 2019 | United States | Scores from the 5 sleep measures were summed<br><br>range of 0 to 5, with higher scores indicating greater sleep health | mix of self-report and objective | 738 | 55.4 (16.2) | African American: 98% | Male: 22% | 5 | Satisfaction/Quality<br>Timing<br>Duration<br>Efficiency<br>Regularity |
| Dong | 2019 | United States | An aggregate measure of sleep health was calculated by summing the number of dimensions of "good" sleep health and classified into 6 categories<br><br>the scale ranges from 0 to 6 with higher scores indicating good sleep health | self-report only | 176 | 14.77 (1.84) | Hispanic/ Latino n=27 (15%)<br><br>White n=114 (65%); African American/black n=12 (7%), American Indian or Alaska Native n=0 (0%), Asian n=18 (10%); Native Hawaiian/other Pacific Islander n=2 (1%); Mixed race n=30 (7%) | female n = 102 (58%) | 6 | Satisfaction/Quality<br>Timing<br>Alertness/Sleepiness<br>Duration<br>Efficiency<br>Regularity |
| Dong | 2023 | United States | The sleep health composite was calculated by summing the 6 binary variables for individual dimensions<br><br>the scale ranges from 0-6 with higher scores indicating good sleep health | mix of self-report and objective | 142 | 14.03 (1.37, 12-16) | 100% American Indian/ Alaska Native (AI/AN)<br>AI/AN: 128 (90%)<br>Hispanic/ Latino(a) 73 (51%)<br>Asian American/Pacif | Male: 58 (41%)<br>Female: 84 (59%) | 6 | Satisfaction/Quality<br>Timing<br>Alertness/Sleepiness<br>Duration<br>Efficiency<br>Regularity |

Supplementary File 4: Data Extraction

| | | | | | | | | | | |
|---|---|---|---|---|---|---|---|---|---|---|
| | | | | | | | ic Islander 13 (9%)<br>Asian 3 (2%)<br>Black/African American 13 (9%)<br>White American 18 (17%)<br>Other 3 (2%) | | | |
| Ensrud | 2020 | United States | A multidimensional measure of poor sleep health (range 0-5) was calculated by summing the number of dimensions with poor scores and categorized as 0, 1, 2, or at least 3 impairments in sleep health dimensions<br><br>The scale ranges from 0-5 higher number indicating poorer sleep health | self-report only | 1459 | 83.6 (3.90%) | African American n=173 (11.9%) | 100% Female | 5 | Satisfaction/Quality<br>Latency<br>Timing<br>Alertness/Sleepiness<br>Duration |
| Furihata | 2017 | United States | An aggregate measure of sleep health was calculated by sum-ming the number of dimensions with "poor" sleep health, and classified into 5 categories: 0, 1, 2, 3, 4, or more<br><br>The scale ranges from 0-5 with higher score indicating poorer sleep health | self-report only | 6485 | 70-74 y: n=328 (5.1%); 75-79 y: n=3063 (47.2%); 80-84 y: n=2041 (31.5%); 85+: n=1053 (16.2%) | Caucasian n=5919 (91.3%); African American n=545 (8.4%); Other n=21 (0.3%) | 100% Female | 5 | Satisfaction/Quality<br>Timing<br>Alertness/Sleepiness<br>Duration<br>Latency |
| Furihata | 2020 | Japan | A composite sleep health score was calculated by summing the number of "poor" sleep health dimensions and classified into five categories: 0, 1, 2, 3, 4 or more.<br><br>Classified into five categories: 0, 1, 2, 3, 4 or more | self-report only | 2482 | 20-29: n = 1350 (54.4%); 30-39: n=688 (27.7%); 40+: n=444 (17.9%) | +Not provided | 100% Female | 5 | Satisfaction/Quality<br>Timing<br>Alertness/Sleepiness<br>Duration<br>Sleep Disorders |
| Furihata | 2024 | Japan | A composite sleep health score was calculated by summing the number of "poor" sleep health dimensions and classified into four categories: 0, 1, 2, 3, or more | self-report only | 1724 | 52.7 +/- 10.7 years | +Not provided | 92.1% male | 5 | Satisfaction/Quality<br>Efficiency<br>Timing<br>Alertness/Sleepiness<br>Duration |

Supplementary File 4: Data Extraction

| | | | | | | | | | | |
|---|---|---|---|---|---|---|---|---|---|---|
| | | | The scale ranges from 0-5 with a higher score indicating poorer sleep health | | | | | | | |
| Griggs | 2022 | United States | The Sleep Health Composite was coded as 1 = good and 0 = poor<br><br>Score ranged from 0 to 5, and higher scores indicating better sleep health | self-report only | 75 | 21.47 (SD 2.056) | Non-Hispanic White n=65 (86.7%) | female n=56 (74.7%) | 5 | Satisfaction/Quality<br>Timing<br>Alertness/Sleepiness<br>Duration<br>Efficiency |
| Harvey | 2021 | United States | each dimension was dichotomized such that 1 = good; 0 = poor<br><br>The scale ranges from 0-6 with higher scores indicate better sleep health. | self-report only | UC-DT n=60, TranS-C + UC (n=61) | UC-DT 45.45 (SD 13.25); TransS-C+UC 47.97 (SD 11.51) | UC-DT<br>Hispanic or Latino n=9 (15%)<br>Not Hispanic or Latino n=51 (85%)<br>Missing (no data entered)<br>TransS-C+UC<br>Hispanic or Latino n=10 (16.39%)<br>Not Hispanic or Latino n=50 (81.97%)<br>Missing n=1 (1.64%)<br>UC -DT<br>White n=21 (35%)<br>African American/Black n=26 (43.3%)<br>American Indian or Alaskan Native n=4 (6.67%)<br>Asian n=5 (8.33%)<br>Native Hawaiian/Other Pacific Islander n=2 (3.33%)<br>Missing n=2 (3.33%) | UC-DT female n=33 (55%); TransS-C+UC female n=30 (49.18%) | 6 | Satisfaction/Quality<br>Timing<br>Alertness/Sleepiness<br>Duration<br>Efficiency<br>Regularity |

| | | | | | | | | | | |
|---|---|---|---|---|---|---|---|---|---|---|
| | | | | | | | TransS-C+UC White n=25 (40.98%) African American/Black n=26 (42.62%) American Indian or Alaskan Native n=4 (6.56%) Asian n=2 (3.28%) Native Hawaiian/Other Pacific Islander n=1 (1.64%) Missing n=2 (3.33%) | | | |
| Hawkins | 2023 | United States | Calculated global sleep health by taking the arithmetic sum of the number of good sleep health indicators.<br><br>The score ranged from 0 to 7, with higher values indicating better sleep health. | self-report only | 1016 | 52 (38,63) | White n=787 (77%); Black n=149 (15%); Asian n=29 (2.9%); Pacific Islander, American Indian, or other races n=17 (1.7%); 2 or More Races n=34 (3.3%) | (Gender) Women n=791 (78%); Men n=225(22%) | 7 | Satisfaction/Quality<br>Timing<br>Alertness/Sleepiness<br>Continuity (2)<br>Duration<br>Regularity |
| Hawkins | 2023 | United States | composite sleep score was based on an aggregate of "healthy" sleep indicators<br><br>The composite sleep score ranged from 0 to 5, with higher scores suggesting better sleep health | objective only | 745 | 27 (23,31) | Non-Hispanic white n=478 (64%); Non-Hispanic black n=88 (12%); Hispanic n=112 (15%); All other races/ethnicities n=67(9%) | 100% Female | 5 | Timing<br>Napping<br>Duration<br>Efficiency<br>Regularity |
| Huang | 2021 | Australia<br><br>(UK Biobank Data) | sum of healthy sleep characteristics<br><br>healthy: ≥ 4; intermediate: 2–3; poor: ≤ 1) | self-report only | 38601 | 55.7 (7.6 SD) | +Not provided | female n=19,713 (51%) | 5 | Alertness/Sleepiness<br>Duration<br>Sleep Disorders (2)<br>Chronotype |

Supplementary File 4: Data Extraction

| | | | | | | | | | | |
|---|---|---|---|---|---|---|---|---|---|---|
| Kline | 2021 | United States | Each dimension was dichotomized as “good” or “poor” on the basis of clinically and/or scientifically relevant rationales. The number of “good” dimensions were summed to provide a composite measure of sleep health.<br><br>The scale ranges from 0-6 with higher values indicating better sleep health | mix of self-report and objective | 125 | 50.3 ± 10.6 | White n=101 (80.8%), Other n=24 (19.2%) | Female n=114 (91.2%) | 6 | Satisfaction/Quality<br>Timing<br>Alertness/Sleepiness<br>Duration<br>Efficiency<br>Regularity |
| Lee | 2020 | United States | Summed the scores of these binary indicators (0 = not having the condition, 1 = having the condition) to construct a composite score of sleep health<br><br>Higher scores represented poorer sleep health (range 0-5) | mix of self-report and objective | 2562 | 76.19 ± 5.49 | +Not provided | 100% Male | 5 | Satisfaction/Quality<br>Alertness/Sleepiness<br>Continuity<br>Napping<br>Duration |
| Lee | 2021 | United States | Then summed binary values across the six indicators.<br><br>The total composite sleep health score ranged from 0 to 6. Higher scores indicated more sleep health problems. | mix of self-report and objective | 441 | 56.85 (11.44) | White n=297 (67%); Black or African American n=133, 30%; Some other race n=11, 3%. | male n=175, 40% | 6 | Satisfaction/Quality<br>Timing<br>Alertness/Sleepiness<br>Duration<br>Efficiency<br>Regularity |
| Lee | 2024 | United States | For each sleep dimension, suboptimal sleep was coded as 1 (vs 0 = optimal sleep). These binary indicators were summed, so that possible scores ranged from 0 to 5 for the sleep health composite. Higher numbers indicated a greater number of sleep health problems<br><br>Higher values indicated a greater number of sleep health problems (Range = 0-5) | self-report only | Baseline (n=5140 ), Change in sleep health n=2991 | Baseline (mean, SD) 56.4 (12.3); Change 63.4 (11) | Non-hispanic White Baseline n=4034 (78%), Change 2330 (78.4%); Non-hispanic black Baseline n=726 (14.2%), Change n=370 (12.5%). Hispanic and all other races Baseline n=359 (7%), Change 272 (.1%). | Female Baseline n2824 (54.9%), Change n=1700 56.8%. Male Baseline n=2316 (45.1%), Change n=1201 (43.2%). | 5 | Satisfaction/Quality<br>Alertness/Sleepiness<br>Duration<br>Efficiency<br>Regularity |
| Madrid-Valero | 2021 | United Kingdom | Coding assigned to responses ( 1-4/5) and then answers summed to create composite. | self-report only | 19457 | mean 57.1 (14.1), range 18-99 | +Not provided | 74.2% female, 24.3% male, 0.5% non-binary, 0.4% preferred not to say, and | 5 | Satisfaction/Quality<br>Latency<br>Continuity (2)<br>Duration |

Supplementary File 4: Data Extraction

| | | | | | | | | | | |
|---|---|---|---|---|---|---|---|---|---|---|
| | | | Overall, scores could range from 4 to 22, where higher scores represent poorer sleep quality. | | | | | 0.5% preferred to self-describe | | |
| Makarem | 2022 | United States | The individual sleep dimensions' scores were summed to create an overall MDSH score ranging from 0–5, such that higher scores were indicative of more favorable sleep health. Further, the score was categorized as 0–1, 2–3, and 4–5 that captured poor, moderate, and ideal sleep health, respectively.<br><br>score ranging from 0–5, such that higher scores were indicative of more favorable sleep health. | self-report only | 4555 | 49 +/- 18 years (range: 20-80 years) | 63% non-Hispanic White<br>11% Non-Hispanic Black<br>16% Hispanic<br>6% Non-Hispanic Asian<br>4.6% Other Race/Multi-Racial | Women: 51.1%<br>Men: 48.9% | 5 | Latency<br>Alertness/Sleepiness<br>Duration<br>Sleep Disorders<br>Regularity |
| Polanka | 2023 | United States | An aggregate measure of sleep health was calculated by summing the number of dimensions with "good" sleep health and classified into 6 domains<br><br>The scale ranges from 0-6 with higher scores indicating better sleep health | mix of self-report and objective | 49 | 64.8 (11.4) | N=20 (40.8%) Black race | Female n=36, 73.5% | 6 | Satisfaction/Quality<br>Timing<br>Alertness/Sleepiness<br>Duration<br>Efficiency<br>Regularity |
| Sampasa-Kanyinga | 2022 | United Kingdom | total number of health sleep characteristics<br><br>the overall sleep pattern was categorized into three groups based on total number of healthy sleep characteristics (healthy: ≥4, intermediate: 2-3, poor: ≤1) | self-report only | 31361 | 56.1 (7.5) | +Not provided | female n(%) 16,281 (52)<br>male 15,080 (48) | 5 | Alertness/Sleepiness<br>Duration<br>Sleep Disorders (2)<br>Chronotype |
| Savin | 2024 | United States | To produce an overall sleep health score, each component was dichotomized with 0 indicating poor sleep health and 1 indicating good sleep health and then summed<br><br>the scale ranges from 0-6 with higher scores indicating better sleep health | mix of self-report and objective | 342 | 43.9 (11.8) | Mexican heritage 341 (99.7%) | female n=226 (66.1%) | 6 | Satisfaction/Quality<br>Timing<br>Alertness/Sleepiness<br>Duration<br>Efficiency<br>Regularity |

Supplementary File 4: Data Extraction

| | | | | | | | | | | |
|---|---|---|---|---|---|---|---|---|---|---|
| Schiel | 2022 | United Kingdom | an aggregate measure of sleep health was calculated by summing the number of dimensions with "good" sleep health and classified into 5 domains<br><br>the scale ranges from 0-5 with higher scores indicating better sleep health | self-report only | 25,758 | 62.9 ± 7.4 years | +Not provided | women n=13993 (54.3%), men n=11765 (45.7%) | 5 | Alertness/Sleepiness<br>Duration<br>Sleep Disorders (2)<br>Chronotype |
| Thomas | 2024 | United States | An aggregate measure of sleep health was calculated by summing the number of dimensions with "good" sleep health and classified into four domains<br><br>the scale ranges from 0-4 with higher scores indicating better sleep health | objective only | 13 | 54.80 (5.2) | White: 13 (100%) | Female: 10 (76.9%) | 5 | Timing<br>Alertness/Sleepiness<br>Duration<br>Efficiency<br>Regularity |
| Tighe | 2021 | United States | Summed scores across sleep health domains to yield a composite multidimensional sleep health score<br><br>(range 0-6), with higher scores indicating better sleep health. | mix of self-report and objective | 158 | 65-85, average 71.8 (5.4) | White or non-Hispanic, n=132, 83.5%<br>Non-white, n=26, 16.5% | Female, n=82, 51.0% | 6 | Satisfaction/Quality<br>Timing<br>Alertness/Sleepiness<br>Duration<br>Efficiency<br>Regularity |
| Turner | 2024 | Australia | The sum of the dichotomized extreme domains for each person is used as a sleep heath composite score<br><br>The scale ranges from 0-7 with higher scores indicating worse sleep health. The cut off ranges were calculated for the CS group and were then used for each of the population groups | mix of self-report and objective | 111 | MS: 50.49 ± 9.63 years HD: 41.47 ± 11.90 years CS: 43.69 ± 13.98 years | +Not provided | MS Female: 81% (n = 35)<br>HD Female: 63% (n = 12)<br>CS Female: 65% (n = 32) | 7 | Satisfaction/Quality<br>Timing<br>Alertness/Sleepiness<br>Continuity<br>Duration<br>Regularity<br>Rhythmicity |
| Wallace | 2018 | United States | Aggregate measure of extreme sleep characteristics calculated by summing number of dimensions with extreme characteristics. Considered continuously in one model and categorically (0, 1, 2, 3, ≥4 extreme characteristics) in another. | mix of self-report and objective | 2887 | 76.3 (5.5), range from 67-96 | Caucasian n=2,595 (89.9%) | 100% Men | 7 | Satisfaction/Quality<br>Timing<br>Alertness/Sleepiness<br>Continuity<br>Duration<br>Regularity<br>Rhythmicity |

Supplementary File 4: Data Extraction

| | | | | | | | | | | |
|---|---|---|---|---|---|---|---|---|---|---|
| | | | The scale ranges from 0-7 with higher scores indicating more poor sleep health | | | | | | | |
| Whibley | 2021 | United States | The number of sleep domains in extreme categories were summed for a composite sleep score.<br><br>0-7, higher scores indicating more extreme sleep domain scores | mix of self-report and objective | 49 | 47.6 (SD 10.0) | +Not provided | 61% female | 7 | Satisfaction/Quality<br>Timing<br>Alertness/Sleepiness<br>Continuity<br>Duration<br>Regularity<br>Rhythmicity |
| Woo | 2024 | United States | Poor sleep health was coded as 0, good sleep health as 1. Scores were summed across dimensions to create a composite.<br><br>Sleep health score ranging from 0 to 6, with higher scores indicating better sleep health. | mix of self-report and objective | 257 | 62.5 ± 10.6 | 100% African American | 72% female | 6 | Satisfaction/Quality<br>Timing<br>Alertness/Sleepiness<br>Duration<br>Efficiency<br>Regularity |
| Xing | 2024 | United States | Based on the sleep composite score, which is the sum of individual sleep scores, participants were divided into four groups: Q1 (less than or equal to 4), Q2 (5-6), Q3 (7-9) and Q4 (greater than or equal to 10).<br><br>The scale ranges from 0-22 with higher scores indicating poor sleep health | objective only | 5706 | 40-90, average 63.2 | white 4843 (84.9%); black 505 (84.9%); other 358 (6.3%) | 47.6% (n=2715) male | 6 | Sleep Characteristics(4)<br>Sleep Disorders<br>Sleep Efficiency |
| Yoo | 2023 | United States | Each dimension was scored as 0 (poor) or 1 (good) with a higher score reflecting better sleep health.<br><br>Defined good overall sleep health as a score of ≥ 3, corresponding to about the 25th percentile | Self-Report Only | 98 | SHS <3: 34.9 ± 11.6; SHS ≥3: 35.1 ± 12.2; overall mean age: 35 ± 12 years | SHS <3: White 12 (80), Black 0, Asian 1 (6.7), multiple other 2 (13.3); SHS > or =3: white 69 (83.1), black 4 (4.8), Asian 3. (3.6), multiple/other 7 (8.4) | 74 were premenopausal women, 12 were menopausal women, and 12 were men; 87.8% female | 6 | Satisfaction/Quality<br>Timing<br>Alertness/Sleepiness<br>Duration<br>Efficiency<br>Regularity |
| Yoo | 2023 | United States | Each dimension was scored as 0 (poor) or 1 (good) with a higher score reflecting better sleep health. | Objective Only | 98 | SHS <3: 34.9 ± 11.6; SHS ≥3: 35.1 ± 12.2; | SHS <3: White 12 (80), Black 0, Asian 1 (6.7), multiple other | 74 were premenopausal women, 12 were menopausal | 4 | Timing<br>Duration<br>Efficiency<br>Regularity |

| | | | | | | | | | | |
|---|---|---|---|---|---|---|---|---|---|---|
| | | | Defined good overall sleep health as a score of ≥ 3, corresponding to about the 25th percentile | | | overall mean age: 35 ± 12 years | 2 (13.3); SHS > or =3: white 69 (83.1), black 4 (4.8), Asian 3. (3.6), multiple/other 7 (8.4) | women, and 12 were men; 87.8% female | | |
| Yu | 2024 | China | Composite Index = –3.914221 + PSQI * 0.0555522 +RBDSQ*0.7287495 + ESS * 0.0660809 | self-report only | 816 | In the Whole pop-ulation, Training cohort, and Validation cohort, the ages of the patients were 67.14 ± 7.61, 67.12 ± 7.58,and 67.21 ± 7.68 years old, respectively | +Not provided | The pro-portion of males was 37.08%, 37.52%, and 36.27%,respectively | 4 | Satisfaction/Quality Alertness/Sleepiness Sleep Disorders (2) |

+Not provided indicates instances where the original article did not specify
